# Antiphospholipid antibodies exacerbate damage following oxygen deprivation-reperfusion injury in an *in vivo* model for stroke and in *ex vivo* blood derived endothelial cells

**DOI:** 10.1101/2024.01.04.24300876

**Authors:** Charis Pericleous, Daniel J. Stuckey, Robert T. Maughan, Koralia Paschalaki, Lida Kabir, Lauren T. Bourke, Rohan Willis, Anisur Rahman, Anna M. Randi, Deepa J. Arachchillage, Mark Lythgoe, Ian P. Giles, Justin C. Mason, Yiannis Ioannou

## Abstract

**Background:** Prothrombotic antiphospholipid antibodies (aPL) found in patients with antiphospholipid syndrome (APS) are a recognised risk factor for ischemic stroke. However, it is unclear if aPL cause injury post thrombolysis leading to worse outcomes. We investigated whether aPL exacerbate reperfusion injury and sought to translate our findings in endothelial colony forming cells (ECFC) isolated from patients with APS.

**Methods:** Transient ischemic stroke was induced in adult rats injected with serum-derived IgG from patients with APS (APS-IgG, containing aPL) or healthy controls (HC-IgG). Infarct size and intracellular signalling processes involved in ischemia-reperfusion injury were determined post reperfusion. *In vitro*, human umbilical vein endothelial cells (HUVEC) treated with IgG, as well as APS and HC ECFC, were exposed to hypoxia (0.1% O_2_). Cell death and relevant signalling mechanisms were assessed following reperfusion and compared to matched normoxic cultures.

**Results:** *In vivo*, APS-IgG induced >2-fold larger infarcts and lower levels of active phosphorylated Akt, a key pro-survival kinase, compared to HC-IgG. *In vitro*, aPL-mediated cell death and suppression of Akt phosphorylation was confirmed in HUVEC exposed to IgG and hypoxia-reperfusion. Consistent with these findings, higher rates of cell death and reduced Akt phosphorylation following reperfusion were observed in *ex vivo* APS ECFC compared to HC ECFC. Treatment with the immunomodulating agent hydroxychloroquine ameliorated ECFC death and this effect was more pronounced in APS-derived cells.

**Conclusion:** Patient-derived IgG aPL exacerbate cell death following reperfusion in a novel *in vivo* stroke model for APS, as well as *in vitro* HUVEC cultures. These observations are mimicked in *ex vivo* APS ECFC. Our findings describe a novel pathogenic role for aPL in mediating tissue injury in addition to their known thrombogenic properties and indicate potential for pharmacological intervention.

## Introduction

Stroke remains a significant cause of morbidity and mortality worldwide. Ischemic stroke accounts for >80% of cases and 10-15% of those occur in young adults.^1^ Approximately 10-20% of patients under 50 years old with stroke have antiphospholipid syndrome (APS), the most common identifiable cause of acquired hypercoagulability in the general population.^2,3^ APS is clinically defined by thrombotic events and/or obstetric complications with persistent presence of circulating antiphospholipid autoantibodies (aPL).^4^ aPL positivity is a strong risk factor for stroke, particularly in young patients and in the absence of classical cardiovascular risk factors (e.g. hyperlipidemia, hypertension, smoking, metabolic disease). Risk of recurrent thrombosis is also elevated in aPL-positive young patients. A prospective study of >1800 patients between 18-45 years old and previous ischemic stroke identified aPL as an independent predictor of long-term recurrent vascular events after first stroke using a composite endpoint of stroke, transient ischemic attack, myocardial infarction (MI) or other arterial events.^5^

Pathogenic aPL are primarily raised against plasma cofactors that bind phospholipids; more recently pathogenic aPL that directly target phospholipids have also been described.^6^ To date the plasma protein beta-2 glycoprotein I (β2GPI) has been identified as the most important autoantigen. The ability of β2GPI to bind cell membrane receptors and phospholipids facilitates aPL priming of endothelium, platelets, monocytes and neutrophils resulting in a proinflammatory and procoagulant phenotype.^7^ Thrombosis in APS is a ‘two-hit’ phenomenon: aPL provide the first thrombophilic ‘hit’ and clotting ensues upon exposure to a second ‘hit’ such as infection or injury. Endothelial injury is central to APS pathogenesis, and existing animal models convincingly demonstrate the prothrombotic nature of aPL and endothelial involvement in carotid, cardiac, mesenteric and femoral vascular territories.^8–14^

Restoration of blood flow with thrombolysis is the first-line treatment for occlusive ischemic events. However, organ reperfusion can exacerbate tissue damage. The extent of reperfusion injury is ultimately determined by the complex interplay between intracellular ‘live-or-die’ signals modulated by the activity of key protein kinases. In cardiac ischemia-reperfusion injury, the role of protein kinase B (PKB/Akt) and p38-mitogen activated protein kinase (MAPK)/extracellular signal-regulated kinase (ERK) signalling are well established and directly relate to myocardial damage.^15^ Similarly, these kinases are implicated in brain ischemia-reperfusion injury and neuronal survival.^16,17^ aPL can regulate all three kinases.^7,18–24^ We previously demonstrated that aPL engage p38-mitogen activated protein kinase (MAPK) to promote cell death in a model of hypoxia-reperfusion injury *in vitro* using primary rat cardiomyocytes; this mechanism was dependent on the interaction between aPL and the immunodominant region of β_2_GPI.^25^ *In vivo,* β2GPI localizes to the liver and uterus but spreads to the brain when mice are challenged with lipopolysaccharide^26^ suggesting that β2GPI localizes to the brain following a pathogenic insult. Following stroke, β2GPI synthesis is upregulated in the liver, localizes to the brain parenchyma and endothelium, and promotes vascular inflammation.^27^ It is therefore plausible that, following ischemia-reperfusion injury and endothelial blood-brain barrier (BBB) breakdown, aPL entering the brain microcirculation can interact with brain tissue via β_2_GPI.

It is unclear whether aPL increase stroke severity or impact long-term outcomes; studies in patients are limited but suggest increased stroke severity and risk of post-stroke disability,^28^ cognitive impairment^29^ and haemorrhagic transformation associated with longer hospitalisation periods and higher mortality.^30^ We hypothesized that aPL disrupt the fine balance between cell survival and death post ischemia and exacerbate stroke damage during reperfusion by targeting essential survival signalling mechanisms. We employed a well-established rat model of transient middle cerebral artery occlusion (MCAO) to design the first *in vivo* model for stroke in APS and translated our findings in patient endothelial colony forming cells (ECFC), a novel source of patient-derived endothelium isolated from venous blood.^31^

## Methods

### Patients and healthy donors

All research subjects signed informed consent according to the Declaration of Helsinki and approved by the appropriate local, regional and/or national Ethics board (University College London, Imperial College London, University of Texas Medical Branch).

A total of 20 patients with APS and 17 healthy controls (HC) were recruited for this study. Polyclonal IgG was isolated from 14 APS and 12 HC; ECFC were isolated from 12 APS and 12 HC donors. Samples from 6 patients and 7 HC were used for both IgG and ECFC isolation. Demographics and clinical information are listed in Table 1. Positive aPL status was recorded from clinical records and confirmed for IgG anti-cardiolipin (aCL) and anti-β2GPI (aβ2GPI) using in-house assays (see Methods: IgG aPL activity).

**Table 1:**
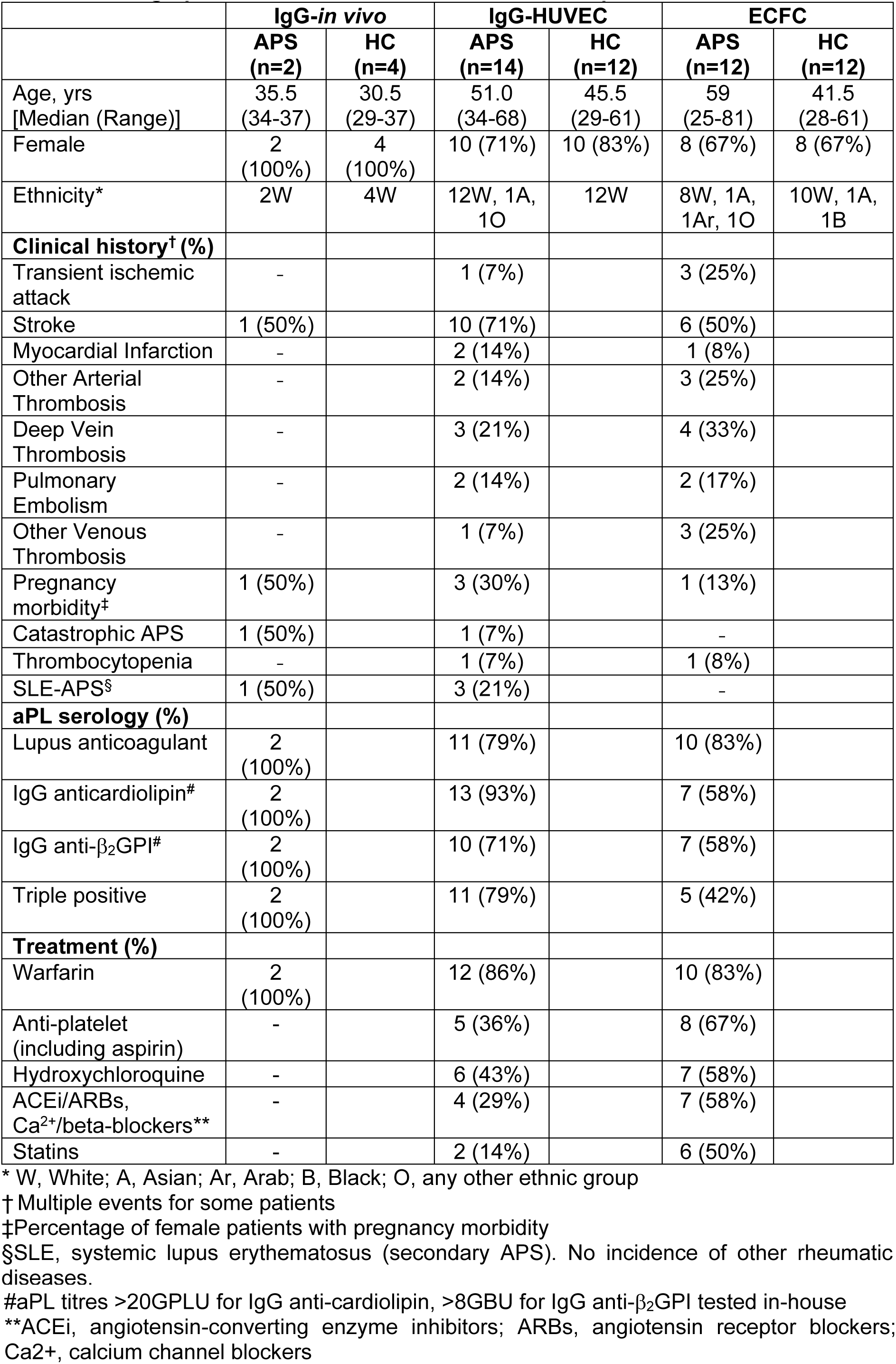
Demographics and clinical characteristics of sampled cohort.

### IgG purification

IgG was purified from human serum by protein G chromatography (ThermoFisher Scientific UK). IgG preparations were concentrated and dialyzed in phosphate buffered saline (PBS) and confirmed to be free of endotoxin (<0.1EU/mL at 1mg/mL concentration of IgG) (EndoLisa, Hyglos Germany).

For *in vivo* experiments, equal amounts of IgG isolated from two APS patients with comparable aPL activity (Table 1) were pooled into a single sample (APS-IgG). A second IgG pool was prepared from four aPL-negative healthy controls (HC-IgG). Both pools were prepared to a concentration of 1mg/mL under sterile conditions. For *in vitro* experiments, IgG samples were tested individually at a concentration of 100μg/mL.

### IgG quantification

Individual and pooled IgG samples were quantified by spectrophotometry and enzyme-linked immunosorbent assay (ELISA).^13,32^ Briefly, 96-well maxisorp plates (Nunc) were coated with an Fc-specific anti-human IgG antibody (Sigma-Merck UK) and blocked to minimize non-specific binding. Serially diluted human IgG was tested in duplicate. Goat anti-human horseradish peroxidase (HRP)-conjugated IgG (Sigma-Merck), teramethybenzidine (TMB) substrate and 0.1M sulphuric acid (KPL Diagnostics, Insight Biotechnology UK) were employed and optical density read at 450nm. IgG concentrations were calculated from a standard curve generated by serially diluting commercially sourced IgG from 100ng/mL to 3.125ng/mL (Sigma-Merck).

Human IgG content was also measured in rodent serum (1:400 dilution)^13,32^ and neat brain tissue lysates (prepared as described below). To confirm assay specificity for human IgG, serum and brain lysates from three rats not injected with human IgG or subjected to MCAO were tested. No signal was detected (data not shown).

### IgG aPL activity

IgG aCL and aβ2GPI activity was measured in purified IgG fractions, human and rat serum using direct ELISA with pre-defined cut offs as previously described.^33^ In brief, 96-well plates were coated with bovine cardiolipin (Sigma-Merck) or human β2GPI (Enzyme Research Laboratories UK). Human serum, rodent serum and purified IgG (serially diluted from 500μg/mL) were tested as described.^13,32^ Signal detection was performed as per the total human IgG ELISA. aCL activity was defined as IgG phospholipid units (GPLU) using validated calibrators (Louisville aPL Diagnostics, USA; activity range 16-120 GPLU). For aβ2GPI, activity was defined as IgG β2GPI units (GBU) using in-house calibrators (activity range 3.1-100 GBU). Titres >99^th^ percentile of the activity of n=200 control sera were considered positive (17GPLU; 8GBU).^33^ LA testing and interpretation was performed by the clinical laboratory following ISTH guidelines.^34^

### In vivo studies

Animal studies were approved by the University College London Biological Services Ethical Review Committee and licensed under the UK Home Office regulations and the Guidance for the Operation of Animals (Scientific Procedures) Act 1986 (Home Office, London, United Kingdom). Male Sprague Dawley (SD) rats (Charles River, UK) weighing between 200-220g, receiving a standard diet and water ad libitum were anaesthetized with 2% isoflurane and injected intravenously with 1mg of pooled APS-IgG or HC-IgG (n=5 per group). The trachea and common carotid artery (CCA) were exposed and isolated. Fifteen minutes post IgG injection, a sterile microfilament was introduced into the CCA and positioned to fully occlude the right MCA.^35,36^ After 0.5hr or 1hr ischemia, the microfilament was removed to enable reperfusion. Animals were allowed to recover, provided with soft tissue bedding, unrestricted access to water and food and euthanized 24hr post reperfusion (endpoint).

Two additional groups of rats (n=6 per group) were injected with 1mg of APS-IgG or HC-IgG but not subjected to MCAO surgical procedures. Animals were maintained for 24hr post injection prior to sacrifice.

Immediately following sacrifice, blood was collected by cardiac puncture for serum isolation. Selected organs were examined and collected for downstream analysis as described below. A schematic of our model is shown in Supplementary Figure 1B.

### Infarct size analysis

Brains isolated from rats subjected to MCAO were cut into 1mm slices and stained with tetrazolium chloride (TTC) to discriminate between metabolically active (red) and dead tissue (white). Representative slices from an APS-IgG treated rat are shown in Figure 1C. A total of 12-13 slices per animal were digitally analysed by a technician blinded to the treatment groups. The area (mm^3^) of viable and infarcted regions (Figure 1C, red and white tissue respectively) was measured on both sides of each slice and lesion size calculated as a percentage of total brain area.

**Figure 1.**
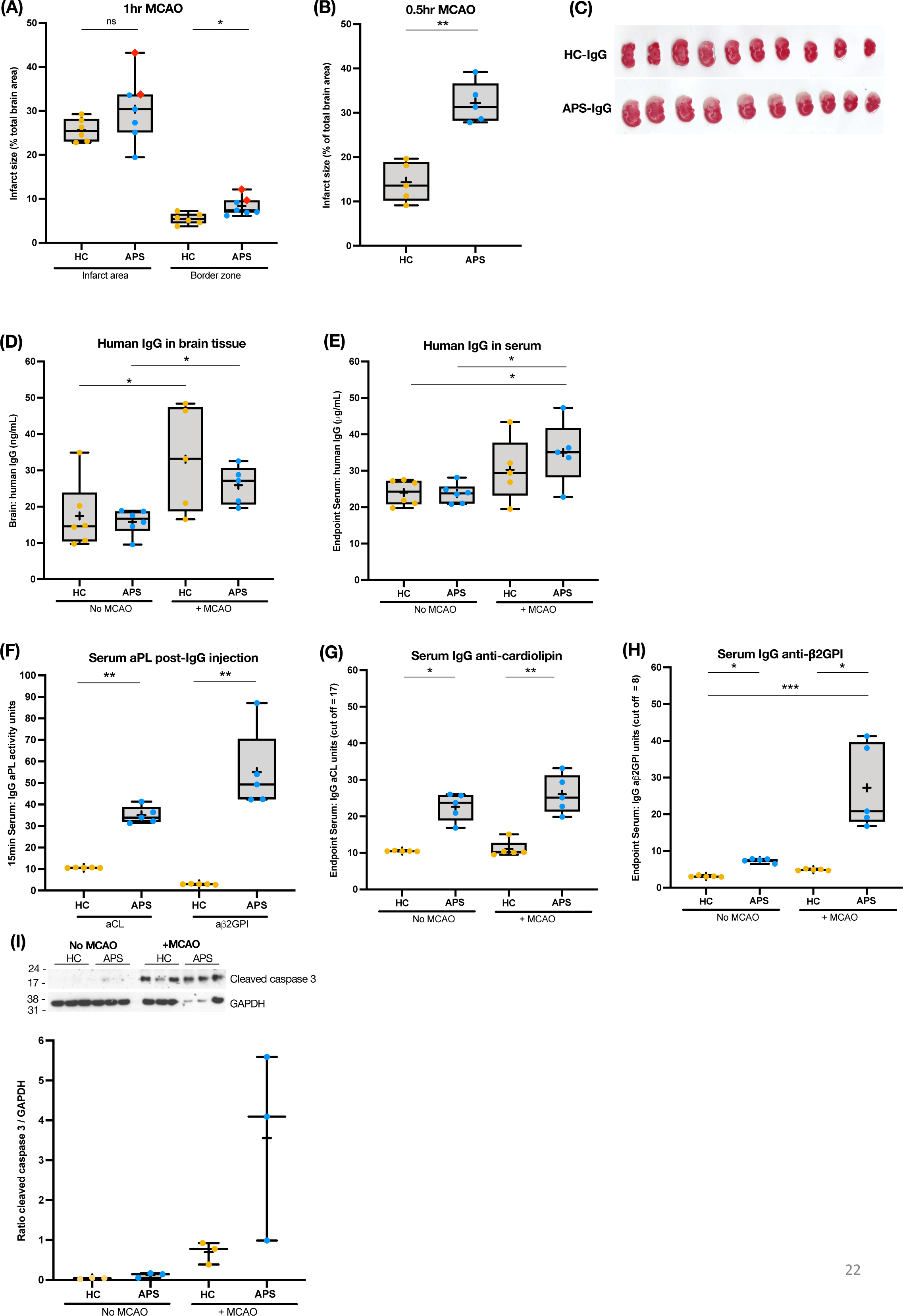
aPL enhance brain injury following ischemia-reperfusion *in vivo*. Rats injected with 1mg human IgG were subjected to transient middle cerebral artery occlusion (MCAO) followed by reperfusion (n=5-8 APS-IgG; n=5-6 HC-IgG). An additional litter of rats were injected with APS-IgG or HC-IgG but not subjected to MCAO (no-MCAO, n=6 per group). **(A)** MCAO was performed for 1hr and reperfusion for 24hr (n=8 APS-IgG; n=6 HC-IgG). A total of 3/8 APS mice died before endpoint (2/3 with completed histology are marked as red diamonds). Analysis was performed after excluding animals that died pre-endpoint. **(B)** MCAO was reduced to 0.5hr followed by reperfusion for 24hr (n=5 per group). Rats administered APS-IgG had significantly larger infarcts at endpoint. **(C)** Example image of brain slices stained with tetrazolium chloride (TTC) to discriminate between metabolically active (red) and dead tissue (white). **(D-E)** Higher levels of human IgG were detected in *(D)* brain lysates and *(E)* serum from MCAO compared to no-MCAO littermates. **(F)** Fifteen minutes post IgG injection and prior to MCAO, rat serum was sampled for IgG aPL activity. Both aCL and anti-β2GPI were detected. **(G-H)** At endpoint, APS-IgG MCAO rat sera were *(G)* aCL and *(H)* anti-β2GPI positive. aCL were also detected in APS-IgG no-MCAO littermates. aPL were undetectable in HC-IgG rats. **(I)** Activation/cleavage of caspase-3 was evident in MCAO brain tissue and was higher in the APS-IgG group (n=3 per group). Normalised to GAPDH. *p<0.05; **p<0.01; ***p<0.001; ****p<0.0001; *ns*, non-significant.

### Endothelial cell culture

Pooled human umbilical vein endothelial cells (HUVEC, Lonza) were cultured in complete endothelial growth medium (EGM2) containing 10% FBS. Experiments were performed at passage 4.

For ECFC generation, peripheral blood mononuclear cells (PBMC) were isolated from whole blood by gradient centrifugation (Ficoll-Paque Plus GE Healthcare). Counted viable PBMC were resuspended in complete EGM2 supplemented with 20% FBS and seeded at ≥0.8x10^6^/ cm^2^ in culture vessels pre-coated with Type I rat tail collagen (BD Bioscience). The media was carefully changed two days post-seeding and repeated every 2-3 days. From five days post-seeding, cultures were routinely screened for the appearance of cobblestone cells that formed dense endothelial colonies over time. Cultures were expanded for 6-8 weeks post-PBMC seeding. Our ECFC isolation protocol is summarized in Supplementary Figure 3A. Representative images of an emerging colony, expanded ECFC and expression of canonical endothelial markers detected by immunofluorescence, are shown in Supplementary Figure 3B. ECFC were confirmed to be of endothelial but not myeloid lineage by flow cytometry (CD31+/CD144+/CD146+/CD45-/CD14-, Supplementary Figure 3C) and ≥98% pure at passage 4. Experiments were performed at passage 4-5.

### Simulated hypoxia-reperfusion *in vitro*

Hypoxia was mimicked by incubating cell cultures in 0.1% O_2_/5% CO_2_ supplemented with nitrogen (H35 hypoxic workstation, Don Whitley Scientific) for 4hrs followed by reperfusion under standard culture conditions (∼20% O_2_) for 0.5hr to 24hrs. Endothelial basal medium (EBM2) in the absence of growth factors or FBS was used during hypoxia. Cells were switched back to complete EGM2 during reperfusion. Where indicated, matched normoxic cultures were kept under standard conditions in EGM2 for the duration of each experiment. Endothelial cultures were routinely visualized to confirm confluency and monolayer robustness throughout the outlined experimental conditions and time points. All measurements were performed in ≥ 2 independent experiments.

HUVEC were pre-incubated with 100μg/mL purified human IgG for 24hr prior to hypoxia-reperfusion simulation. IgG was retained in the culture medium at all times.

ECFC were cultured in the absence of exogenous IgG. For targeted experiments with hydroxychloroquine (HCQ, 10μg/mL), ECFC were pre-treated with the drug for 24hr prior to hypoxia-reperfusion simulation. HCQ was retained in the culture medium at all times.

### Protein extraction

Small brain slices from the posterior part of the brain were collected from all animals (MCAO and no MCAO) and snap frozen in liquid nitrogen. The kidney, liver, spleen, left carotid artery and aorta were also snap frozen. Tissues were ground to a fine powder using a mortar and pestle placed on dry ice. Approximately 150-200μg of ground tissue was mixed with RIPA (ThermoFisher) or cell lysis buffer (Cell Signaling Technologies UK) containing protease and phosphatase inhibitors (Roche) and incubated on ice for 10min. Lysates were passed through a glass homogeniser and incubated on ice for a further 15min, then spun at 17000g for 10min to pellet cell debris. Total protein in collected supernatants was quantified by bicinchoninic acid (BCA) assay (ThermoFisher) and stored at -20°C.

HUVEC and ECFC were placed on ice and washed twice with HBSS, incubated for 10min with lysis buffer and manually scraped off the culture vessel. Lysates were spun, supernatant collected and quantified as above.

### Immunoblotting

10μg protein lysate was mixed with loading buffer containing SDS and dithiothreitol (DTT), boiled at 95°C for 5mins, separated by electrophoresis (4-12% Bolt Bis-Tris pre-cast gels, ThermoFisher) and transferred onto polyvinylidene difluoride (PDVF) membrane (GE Healthcare UK). Membranes were incubated with primary antibodies followed by species-specific HRP-conjugated or fluorescently labelled secondary antibodies (Supplementary Table 1). Proteins were visualized by chemiluminescence after substrate addition (ECL, GE Healthcare) or by fluorescence (LICOR Odyssey imaging system). Molecular target levels were normalised by calculating the ratio between phosphorylated and total target protein or phosphorylated protein and housekeeping/loading controls.

### Immunoprecipitation

100μg tissue lysate in 200μL lysis buffer was mixed with 5μL of Akt antibody and incubated with gentle rocking overnight at 4°C. Then, 20μL of protein G agarose (ThermoFisher) was added for a further 3hr and the mixture centrifuged at 14000g for 30sec. After five washes with cold lysis buffer, the pellet was resuspended in SDS loading buffer and handled as described above.

### Endothelial cell kinetic apoptosis & necrosis assay

HUVEC or ECFC were seeded at 1x10^4^ per well in 96-well white walled clear bottom culture plates and tested in triplicate under hypoxia-reperfusion and normoxic conditions. Following 2hr reperfusion, apoptosis and necrosis was simultaneously measured in hypoxia-reperfusion and matched normoxic plates by luminescence and fluorescence respectively (RealTime-Glo Annexin V Apoptosis and Necrosis Assay, Promega UK). Luminescence and fluorescence measurements were obtained for 24hr post reperfusion and expressed as fold change of apoptosis or necrosis in hypoxia-reperfusion versus matched normoxic cells. Samples were tested in ≥ 2 independent experiments.

As a positive control for this assay, HUVEC under normoxic conditions were treated for 2hr with 100μM hydrogen peroxide (H_2_O_2_) followed by washout/recovery for 24hr. H_2_O_2_ induced both necrosis and apoptosis (Supplementary Figure 2A). Results are expressed as fold change of apoptosis/necrosis between H_2_O_2_ and untreated HUVEC (n=2 individual HUVEC pools).

### Statistical analysis

Results are presented as (A) Box-and-whisker plots with each data point representing an individual sample. The vertical line inside each plot represents the median value for each data set ± interquartile range (IQR); or (B) line graphs, each line representing an individual sample. Two-tailed unpaired Mann Whitney were used to compare 2 experimental groups and Kruskall-Wallis followed by Dunn’s multiple comparisons tests for >2 groups. Where applicable, paired analysis was performed with equivalent paired non-parametric tests. For correlation analysis, Spearman’s tests were applied. Significance in all cases, p<0.05. Experiments with n≤3 per group were not statistically analysed.

## Results

### APS-IgG increase brain infarct size

For *in vivo* studies, purified IgG was pooled from 2 patients with APS and 4 HC (Table 1). Patients were triple aPL positive in serological assays (LA, high titre IgG aCL, IgG aβ_2_GPI) and all HC were negative. Purified IgG samples were individually tested for aPL activity prior to pooling, confirming high-titre aPL activity in APS-IgG (at 100μg/mL, aCL and aβ_2_GPI activity was >100 units for both samples)^33^ and lack of aPL activity in HC-IgG (tested at up to 500μg/mL; Supplementary Figure 1A).

In the first instance, we performed MCAO for 1hr followed by 24hr reperfusion. Brain infarcts in the APS-IgG group tended to be larger than HC-IgG. The border zone, the most peripheral zone of the affected area and with less distinct pallor than the infarcted core, was also extended (Figure 1A). However, three of eight animals injected with APS-IgG (37.5%) died within 2hr reperfusion. Histology performed on two of three deceased rats revealed infarcts >30% of total brain area (Figure 1A, red diamonds). For the remaining five APS-IgG animals that survived 24hr post reperfusion plus the two histologically analysed animals that died, we observed a series of systemic abnormalities during histological analysis. Specifically, we identified brain clots in 4/7 animals; liver clots in 1/7; dark areas across the heart in 5/7; dark areas in the lungs in 1/7; trouble breathing in 1/7; piloerection in 1/7; and viscous blood in 3/7. Only one animal in the HC-IgG group had dark spots on the liver and viscous blood at endpoint.

The sudden death of a substantial proportion of APS-IgG animals combined with our histological observations suggested that experimental conditions were too harsh in the presence of aPL, and potentially resembled a phenotype closer to the clinical syndrome ‘catastrophic APS.’ This rare form of APS is typified by spontaneous development of multiple microthrombi, consequent multi-organ failure and 40-50% mortality despite aggressive immunosuppression. Therefore, we opted to reduce the duration of MCAO to 0.5hr and maintain reperfusion at 24hr (schematic summary, Supplementary Figure 1B).

Under these conditions, APS-IgG animals had 2.3-fold larger infarcts compared to HC-IgG [infarcted % of total brain area, median (IQR) 31.3% (28.2-36.6) versus 13.6% (10.2-18.9), p=0.008; n=5 per group) (Figure 1B-C)]. We singularly noted dark areas in the kidney and liver in two animals and a third had viscous blood. No other abnormalities or premature death were seen.

For comparison, two additional groups of animals received APS-IgG or HC-IgG but were not subjected to MCAO (no-MCAO); endpoint was defined as 24hr post-IgG transfer. In brain lysates prepared from MCAO and no-MCAO groups, human IgG was more abundant in MCAO compared to no-MCAO tissue, confirming BBB breakdown (Figure 1D). Human IgG levels were similar in APS-IgG and HC-IgG MCAO brain lysates suggesting that any downstream differences can be directly attributed to pathogenic aPL found in APS-IgG.

Similarly, circulating human IgG was detected in both APS-IgG and HC-IgG treated rats. Human IgG levels were higher in MCAO compared to no-MCAO littermates and may point towards IgG retention in the circulation following injury (Figure 1E). In line with this, serum aPL activity was detectable in APS-IgG treated rats 15min post-injection (Figure 1F) and at endpoint (Figure 1G-H). While circulating aCL were equally detectable in both MCAO and no-MCAO APS-IgG littermates (Figure 1G), aβ2GPI levels were pronounced in the APS-IgG MCAO group only (Figure 1H) suggesting greater aβ2GPI retention following injury. All HC-IgG rat sera were aPL negative.

### APS-IgG inhibit pro-survival Akt following brain ischemia-reperfusion injury

Consistent with enhanced infarct size in APS-IgG animals, the apoptotic marker cleaved caspase-3 was elevated in APS-IgG compared to HC-IgG animals following MCAO, and minimal or absent in no-MCAO brain lysates (Figure 1I). We assessed whether aPL disrupts pro-survival mechanisms, key for limiting reperfusion injury and initiating repair. Phosphorylation of the pro-survival kinase Akt (p-Akt) was measured on its two critical activation sites (S473, T308); T308 phosphorylation allows initial Akt activation and S473 phosphorylation further enhances its catalytic activity. In the absence of MCAO, p-Akt S473 levels were similar in APS-IgG and HC-IgG animals (Figure 2A, Supplementary Figure 1C). Following MCAO and as expected, p-Akt S473 levels were higher in HC-IgG treated animals compared to their no-MCAO littermates, suggesting induction of pro-survival signalling. However, elevated p-Akt following MCAO was not observed in APS-IgG animals. Comparison of APS-IgG and HC-IgG MCAO groups revealed strikingly lower phosphorylation at both S473 (Figure 2A, Supplementary Figure 1C) and T308 (Supplementary Figure 1D) in the presence of aPL. Levels of p-Akt S473 negatively correlated with infarct size highlighting the protective role of Akt following ischemia-reperfusion injury (r= -0.9048, p=0.005; Supplementary Figure 1E). Phosphorylated levels of other key kinases involved in reperfusion signalling were similar between APS-IgG and HC-IgG animals following MCAO (p38, ERK1/2, JNK1/2; Supplementary Figure 1F). Taken together, our observations indicate specific inhibition of p-Akt by aPL following oxygen deprivation-reperfusion.

**Figure 2.**
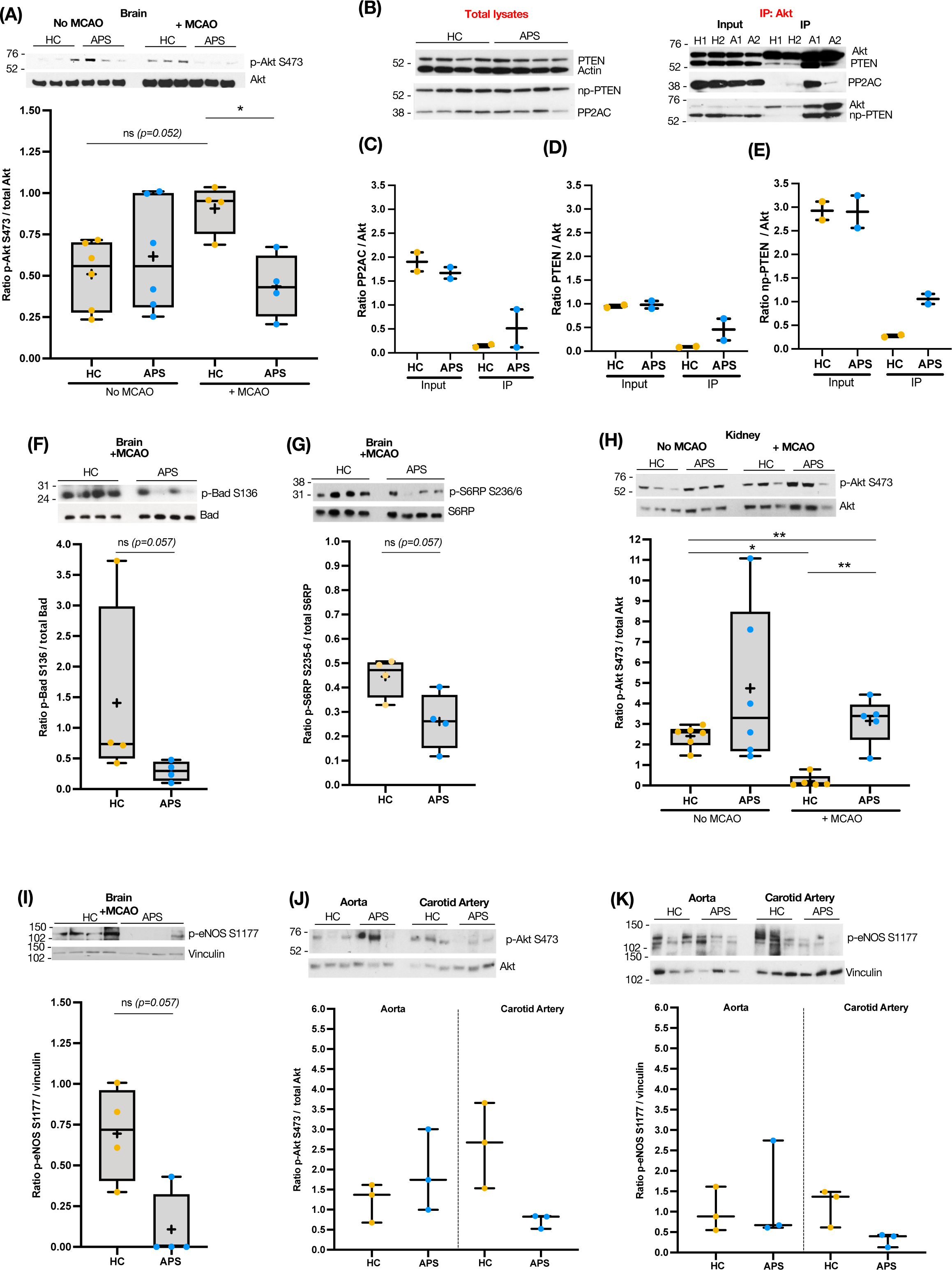
aPL dysregulate the pro-survival kinase Akt. **(A)** Following ischemia-reperfusion injury, HC-IgG animals demonstrated increased phosphorylation of Akt (p-Akt S473) in the brain. In comparison, p-Akt was lower in APS-IgG animals (n=4 MCAO, n=6 no-MCAO per IgG group). **(B-E)** Levels of natural Akt inhibitors PP2A(C, catalytic subunit), PTEN and active PTEN (non-phosphorylated, np-PTEN) were unaltered in the two MCAO groups. Whole lysates *(B, left panel)* or Akt-immunoprecipitated lysates *(B, right panel)* were blotted for *(C)* PP2AC, *(D)* PTEN and *(E)* np-PTEN demonstrating that Akt is bound to active PTEN exclusively in the APS group (n=2 per group). **(F-G)** Analysis of downstream Akt signalling in MCAO brain revealed lower levels of (F) p-Bad S136 and (G) p-S6RP S235/6 in the APS group. **(H)** Higher p-Akt levels were detected in matched MCAO kidney lysates from APS-IgG compared to HC-IgG animals. **(I)** Additional Akt signalling analysis identified lower p-eNOS S1177 levels in brain lysates of APS-IgG compared to HC-IgG animals (n=4 per group). **(J)** p-Akt and **(K)** p-eNOS were lower in APS-IgG carotid artery but not aortic lysates (n=3 per group). For panels A, H representative blots are shown for n=3 animals per MCAO and no-MCAO conditions. Complete blots for all individual lysates tested are provided in Supplementary Figure 1C (brain) and 1G (kidney). Results are normalised as follows: p-Akt, p-Bad, p-S6 to total Akt, Bad, S6 respectively; PP2AC, PTEN, np-PTEN to co-immunoprecipitated total Akt; p-eNOS to vinculin. *p<0.05; **p<0.01; ***p<0.001; ****p<0.0001; *ns*, non-significant.

Previous studies highlight the ability of IgG aPL to interfere with Akt signalling. Canaud *et al* reported aPL-mediated *induction* of p-Akt contributing to endothelial hyperplasia in APS nephropathy^24^ while Sacharidou *et al* demonstrated prothrombotic aPL-mediated *inhibition* via protein phosphatase 2A (PP2A),^37^ one of three molecular Akt inhibitors. We searched for involvement of the molecular Akt inhibitors PP2A, phosphatase and tensin homolog (PTEN) and thioesterase superfamily member 4 (*THEM4*, CTMP) in a limited number of brain lysates from animals subjected to MCAO. Total levels of the PP2A catalytic subunit (PP2AC) and PTEN did not differ between APS-IgG and HC-IgG groups (Figure 2B, left panel). CTMP was not detected. Following targeted Akt immunoprecipitation, evidence favouring PP2A involvement was inconclusive (Figure 2B, right panel and Figure 2C). Instead, an interaction between Akt and PTEN in APS-IgG animals was detected (Figure 2B, right panel and Figure 2D). Akt-bound PTEN was in an activated form as shown by its dephosphorylated state (Figure 2B, right panel and Figure 2E). Since PTEN degradation following stroke may be neuroprotective,^38^ its role and potential manipulation in our model may warrant future exploration.

Next, we studied the catalytic ability of Akt by measuring activation of its downstream substrates in MCAO brain lysates. Phosphorylated levels of the pro-apoptotic BCL2-associated agonist of cell death (p-Bad), which is inhibited by Akt phosphorylation on S136, were reduced in APS-IgG animals (Figure 2F). Consistent with elevated cleaved caspase-3 levels (Figure 1H), a process also regulated by Akt, these observations highlight exacerbated pro-death signalling in the presence of aPL.

Akt supports protein synthesis via phosphorylation of the mammalian target of rapamycin (mTOR) and its downstream effector S6 ribosomal protein (S6RP). Lower p-S6RP levels were detected in the APS-IgG group (Akt target site serine-235/6, Figure 2G). This was an intriguing finding since Canaud *et al* demonstrated elevated p-Akt *and* p-S6RP localized to vascular endothelium in renal biopsies from APS nephropathy patients compared to non-APS disease and healthy controls.^24^ We probed kidney lysates from our animal model and found that, in stark contrast to the brain, p-Akt kidney levels were higher in APS-IgG than HC-IgG following MCAO (Figure 2H, Supplementary Figure 1G). For comparison, we measured p-Akt levels in the liver and spleen but did not detect any differences suggesting that in our model, aPL specifically modulated p-Akt in the brain and kidney (Supplementary Figure 1H).

The APS nephropathy study suggested that aPL activate Akt in the absence of other stimuli.^24^ Upon challenge with exogenous vascular endothelial growth factor (VEGF, a potent Akt inducer), aPL were shown to inhibit activation of the vascular homeostatic enzyme endothelial nitric oxide synthase (eNOS), as demonstrated by reduced phosphorylation of the Akt-regulated S1177 eNOS residue.^37^ We hypothesized that the natural process of Akt induction following ischemia-reperfusion is challenged by aPL and leads to inhibition of p-eNOS, as described in the context of VEGF. We probed brain lysates from our model and noted suppression of p-eNOS in APS-IgG MCAO animals (Figure 2I). In carotid artery specimens collected from the same animals, a similar pattern of p-Akt and p-eNOS inhibition was seen in the APS-IgG group while in the aorta, these molecules were inconsistently affected (Figure 2J, p-Akt and Figure 2K, p-eNOS).

### APS-IgG enhance cell death and inhibit Akt following hypoxia-reperfusion in cultured endothelial cells

Previous aPL-Akt associated studies focused on endothelium,^24,37^ a major aPL target.^7^ Given the importance of endothelial BBB breakdown in stroke, *in vitro* endothelial models were next employed to confirm our findings. Healthy HUVEC treated with APS-IgG or HC-IgG (24hr) were exposed to hypoxia under nutrient restricting conditions (0.1% O_2_, 4hr) followed by reperfusion and nutrient restoration. Cell death was measured in real time for up to 24hr reperfusion and compared to matched cultures kept in normoxic conditions. Sensitivity of the cell death assay was confirmed in HUVEC exposed to H_2_O_2_ (Supplementary Figure 2A) or hypoxia-reperfusion (Supplementary Figure 2B). Comparison of APS-IgG *vs* HC-IgG treated HUVEC responses revealed a non-significant trend for elevated apoptosis following 2hr reperfusion (Figure 3A) and significantly elevated necrosis at all time points (Figure 3B), indicating enhanced endothelial cell death in the presence of aPL.

**Figure 3.**
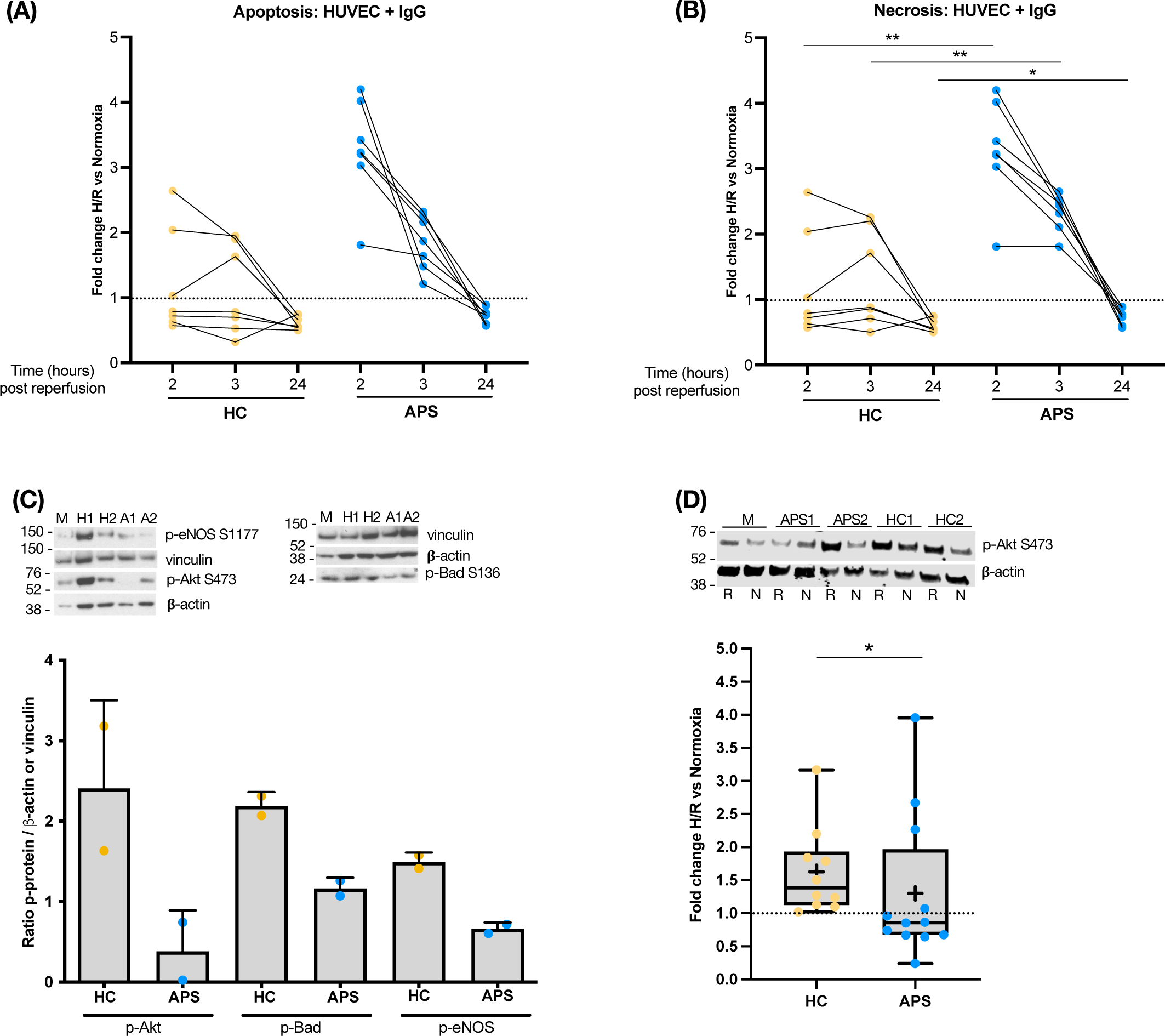
aPL promote endothelial cell death and repress Akt activation following hypoxia-reperfusion injury *in vitro*. HUVEC were incubated with 100μg/mL human IgG for 4hr under hypoxic conditions followed by reperfusion for up to 24hr. Matched cells were treated with IgG and kept under normoxic conditions. **(A-B)** Cell death is expressed as fold change between 2hr, 3hr and 24hr reperfusion *vs* matched normoxic cultures. Higher rates of *(A)* apoptosis and *(B)* necrosis were observed in APS-IgG compared to HC-IgG treated HUVEC following hypoxia-reperfusion (n=7 per group). **(C)** After 2hr reperfusion, cells were harvested for protein analysis. Treatment with APS-IgG resulted in lower p-Akt S473, p-Bad S136 and p-eNOS S1177 levels compared to HC-IgG (n=2 per group). Results are normalised to loading controls: p-Akt/p-Bad to β-actin and p-eNOS to vinculin. **(D)** Extended IgG experiments revealed reduction of p-Akt following reperfusion. Representative immunoblots for two APS-IgG and two HC-IgG treated cultures under hypoxia-reperfusion (R) or normoxia (N), and untreated cells (media alone, M). Immunoblots for the remaining IgG samples tested are provided in Supplementary Figure 2E. Results are shown as fold change of p-Akt levels (normalised to β-actin) at 2hr reperfusion *vs* normoxia (n=12 APS-IgG, n=10 HC-IgG). *p<0.05; **p<0.01; ***p<0.001; ****p<0.0001; *ns*, non-significant.

We sought to establish whether aPL influence endothelial Akt *in vitro*, building on evidence from our *in vivo* observations and from published aPL-endothelial studies.^24,37^ Initially, we tested the same samples used to create pooled IgG for *in vivo* injections (n=2 APS, n=2 HC) and selected phosphorylated targets were measured after 2hr reperfusion. In untreated HUVEC, both p-Akt and p-Bad were elevated following hypoxia-reperfusion compared to normoxia (Supplementary Figure 2C-D). In APS-IgG treated HUVEC, reduced levels of p-Akt, p-Bad and p-eNOS were detected under hypoxia-reperfusion and compared to HC-IgG treated cells (Figure 3C). Reduced p-Akt induction in the presence of aPL, was confirmed in an extended sample set of purified IgG (n=12 APS-IgG, n=10 HC-IgG; Table 1) used to treat matched HUVEC cultures under hypoxia-reperfusion or normoxia (Figure 3D, Supplementary Figure 2E). These results further highlight that aPL disrupts Akt signalling following a context-specific insult, in this case hypoxia-reperfusion injury.

### APS ECFC response to hypoxia/reperfusion mimics *in vivo* observations

Next, we employed *ex vivo* ECFC to study the impact of hypoxia-reperfusion injury on APS endothelium (n=12 donors per group; Table 1). ECFC from patients with systemic disease such as APS, are chronically exposed to circulating pathogenic mediators such as autoantibodies and inflammatory cytokines, ultimately leading to a ‘primed’ phenotype that persists *ex vivo*.^31,39^ Utilising APS ECFC in our study would therefore facilitate extension of our *in vivo* and *in vitro* findings to a biologically relevant source of patient endothelium.

ECFC generation parameters as outlined by ECFC standardization guidelines^40^ including number of endothelial colonies detected, day of first colony detection and day of first passage from P0 to P1 (defined as days-post PBMC seeding), were similar for APS and HC ECFC (Supplementary Figure 3D) suggesting that any functional or molecular differences detected were not due to initial isolation and growth capacity of patient *vs* control cells.

All experiments were performed in parallel matched hypoxia-reperfusion and normoxic cultures. Mirroring the HUVEC-IgG experiments, ECFC were exposed to hypoxia and cell death was tracked in real time for 24hr post reperfusion. Hypoxia-reperfusion promoted cell death in both APS and HC ECFC compared to normoxic cells (Figure 4A-B, Supplementary Figure 4A-B). Both apoptosis (Figure 4A) and necrosis (Figure 4B) were significantly more prominent in APS-ECFC.

**Figure 4.**
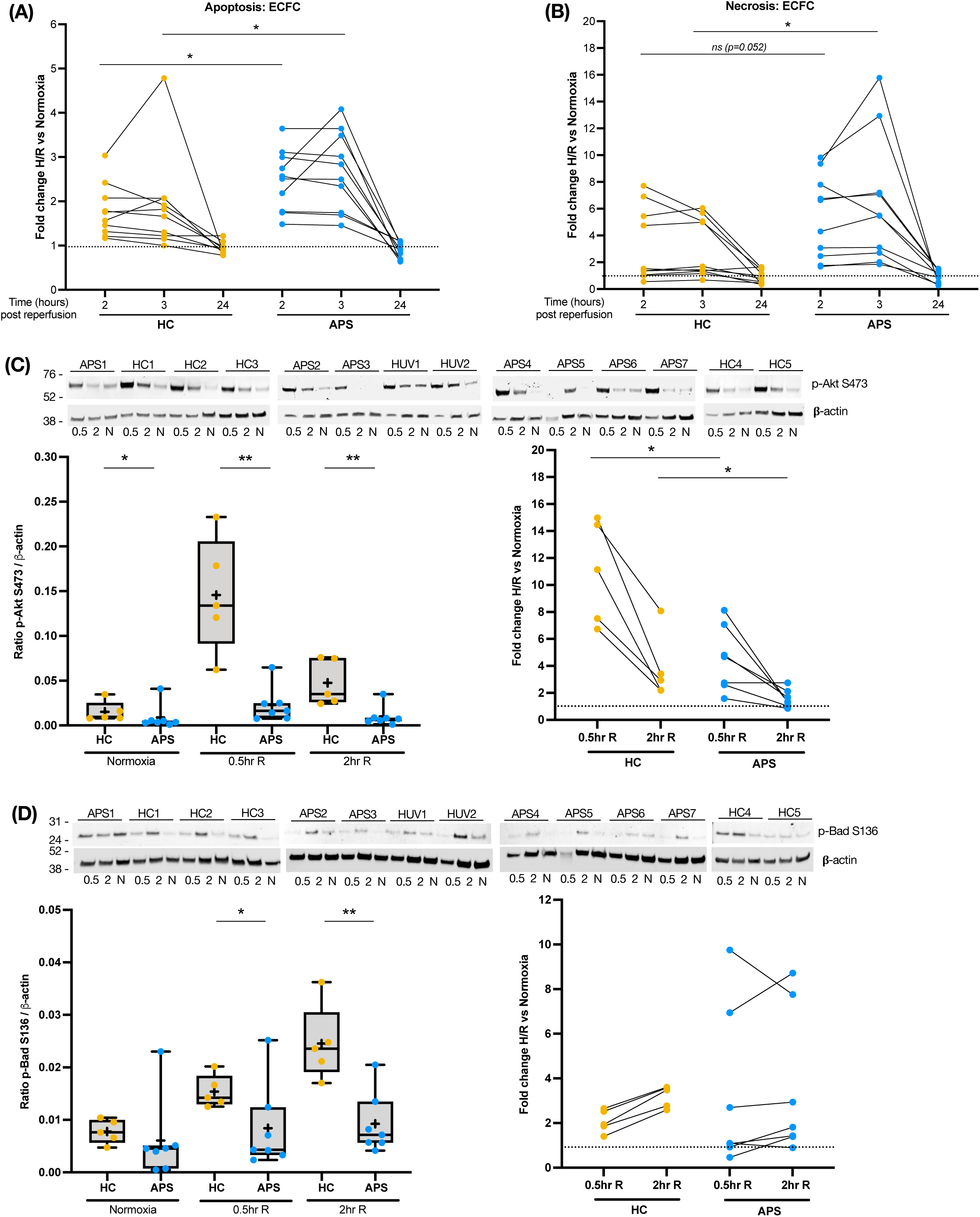
Exacerbated cell death and reduced p-Akt in APS ECFC following hypoxia-reperfusion. Primary ECFC were subjected to hypoxia-reperfusion or normoxia. **(A-B)** Cell death is expressed as fold change between 2hr, 3hr and 24hr reperfusion *vs* matched normoxic cultures. Higher rates of *(A)* apoptosis and *(B)* necrosis were observed in APS compared to HC ECFC following hypoxia-reperfusion (n=10 per group). Luminescence (apoptosis) and fluorescence (necrosis) traces are provided in Supplementary Figure 4A-B. **(C)** Following 0.5hr and 2hr reperfusion, p-Akt levels were lower in APS (n=7) compared to HC ECFC (n=5). Left panel shows p-Akt normalised to β-actin for 0.5hr, 2hr reperfusion (0.5R, 2R) and normoxia (N). Right panel shows fold change of normalised p-Akt between culture conditions. **(D)** Similarly, p-Bad levels were lower in APS compared to HC ECFC. Comparative responses in two untreated HUVEC pools (HUV1, HUV2) are shown for both p-Akt (C) and p-Bad (D); see quantification in Supplementary Figures C-D. Yellow circles, HC; blue circles, APS (n=10 per group). *p<0.05; **p<0.01; ***p<0.001; ****p<0.0001; *ns*, non-significant.

Dysregulation of the p-Akt response following hypoxia-reperfusion was next determined. Induction of p-Akt was observed in both APS and HC ECFC following hypoxia-reperfusion and compared to matched cells kept under normoxia (Supplementary Figure 4C). However, p-Akt levels were robustly lower in APS-ECFC compared to HC-ECFC at both 0.5hr (early) and 2hr (late) post-reperfusion, as well as under normoxic conditions (Figure 4C). Failure to activate Akt resulted in the expected reduction of downstream p-Bad in APS ECFC (Figure 4D, Supplementary Figure 4D). In agreement with the regulatory relationship between Akt and Bad, early p-Akt correlated with late p-Bad (r=0.622, p=0.035; Supplementary Figure 4E). Thus, our *ex vivo* findings in APS and HC ECFC recapitulated our *in vivo* observations and the *in vitro* IgG-HUVEC model.

### Hydroxychloroquine partially protects against hypoxia-reperfusion induced ECFC death

Our group and others reported a protective effect for hydroxychloroquine (HCQ) following brain,^41^ heart^42,43^ and kidney^44^ ischemia-reperfusion injury *in vivo* and in cellular models.^42,44^ We determined whether HCQ could reverse ECFC death following hypoxia-reperfusion. Cells were pre-treated with HCQ for 24hr followed by hypoxia-reperfusion or normoxic culture as described. Under normoxic conditions, HCQ had a variable effect on apoptosis and necrosis (Supplementary Figure 5A-B). Following reperfusion, HCQ suppressed apoptosis (Figure 5A) and necrosis (Figure 5B) in both APS and HC ECFC but yielded a stronger protective effect upon APS ECFC death.

**Figure 5.**
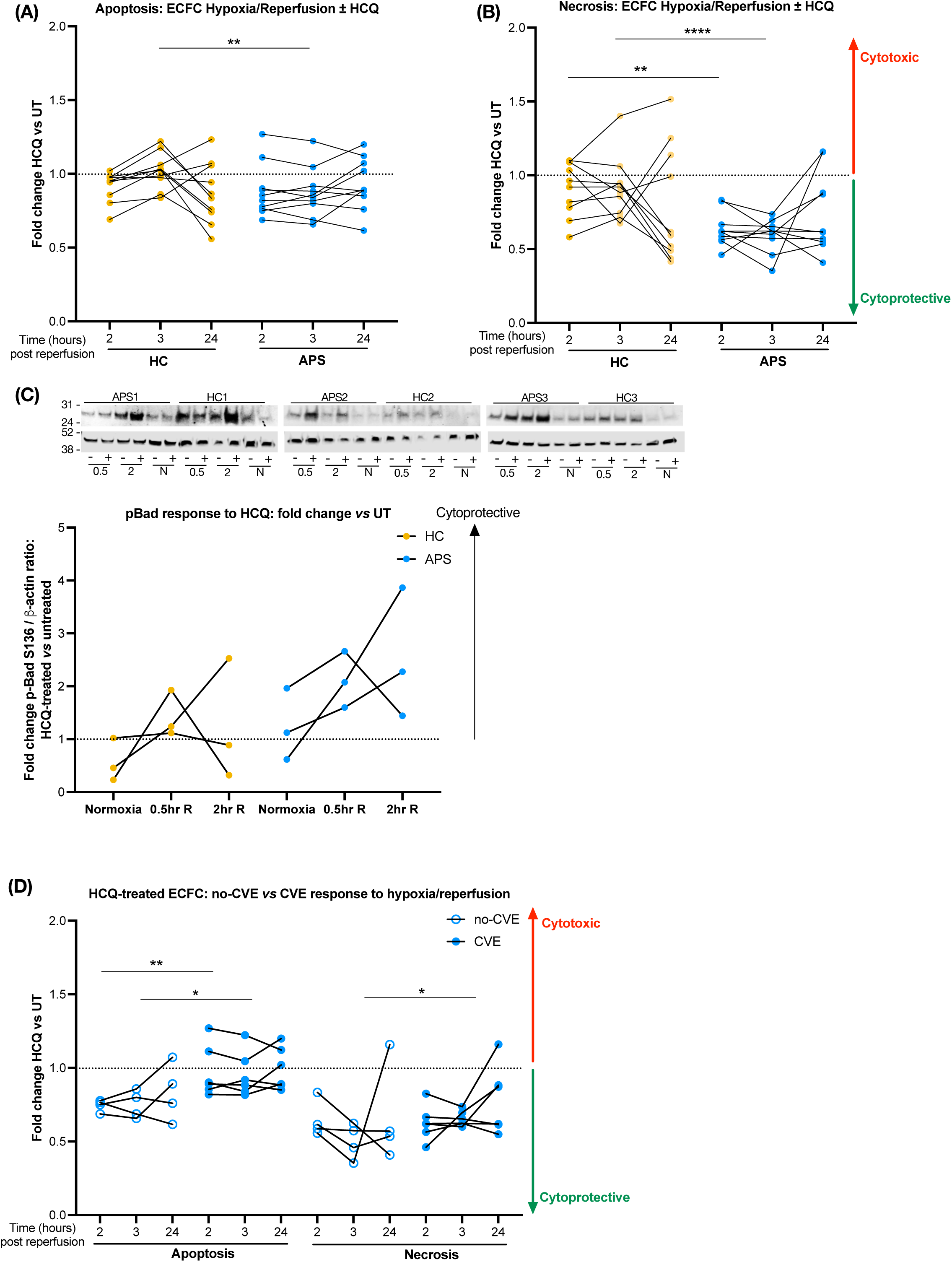
HCQ protects against hypoxia-reperfusion ECFC death *in vitro.* **(A-B)** ECFC were pre-treated with 10μg/mL hydroxychloroquine (HCQ) for 24hr followed by hypoxia-reperfusion or kept under normoxic conditions. Cell death is expressed as fold change between HCQ-treated and untreated cultures. At 2hr and 3hr post reperfusion, HCQ suppressed *(A)* apoptosis and *(B)* necrosis in APS and a proportion of HC ECFC. This protective effect was more prominent in APS ECFC (n=10 per group). **(C)** HCQ promotes elevated p-Bad levels post-reperfusion in both HC and APS ECFC (n=3 per group). **(D)** Separating APS ECFC donors into those with previous cardiovascular events (CVE, n=6) *vs* those without (no-CVE, n=4) revealed greater protection from hypoxia-reperfusion induced cell death in the no-CVE group. Results are expressed as fold change of cell death in HCQ-treated *vs* untreated ECFC. In the absence of HCQ treatment, no obvious difference between cell death rates was observed in CVE *vs* no-CVE groups (Supplementary Figure 5E). *p<0.05; **p<0.01; ***p<0.001; ****p<0.0001; *ns*, non-significant.

We measured p-Akt and p-Bad levels in a small sample set (n=3 per group); p-Akt levels were similar between untreated and HCQ-treated lysates (data not shown) but p-Bad levels appeared to rise following HCQ treatment and hypoxia-reperfusion (Figure 5C, Supplementary Figure 5C). Taken together, our results suggest that HCQ limits cell death during reperfusion in both APS and non-APS cellular sources but its beneficial effects may be enhanced in APS.

In this study, ECFC from thrombotic APS patients with or without a previous history of cardiovascular events (CVE *vs* no-CVE) were used. Comparison of ECFC responses from these two APS subgroups showed equal susceptibility to hypoxia-reperfusion induced cell death (Supplementary Figure 5D). However, *in vitro* treatment with HCQ conferred greater protection in ECFC from no-CVE compared to CVE donors (Figure 5D). Similarly, ECFC responses were compared between patients receiving HCQ compared to those not receiving HCQ at the time of ECFC isolation (HCQ *vs* no-HCQ). No differences in the level of cell death (Supplementary Figure 5E) or response to *in vitro* HCQ treatment was observed (Supplementary Figure 5F).

## Discussion

To our knowledge, we present the first *in vivo* model for aPL-mediated cell death following ischemia-reperfusion injury and identify repression of Akt activity as a critical underlying mechanism. We confirm elevated cell death and reduced p-Akt *in vitro* in HUVEC stimulated with exogenous aPL and subjected to hypoxia-reperfusion. Excitingly, use of *ex vivo* APS ECFC fully recapitulated our *in vivo* observations thus directly extending our findings to a patient-relevant source of endothelium.

Stroke is the most frequent primary arterial thrombotic event in APS and the most frequent recurrent thrombotic event overall (venous and arterial combined).^45–47^ While the relationship between aPL and stroke is well-established, few studies have compared stroke severity and long-term outcomes between APS and non-APS stroke patients, a particularly challenging task as APS strokes occur in younger patients. A study in patients <54 years old identified a correlation between aPL titres, stroke severity on admission and three-month outcomes.^28^ Another study found that cognitive impairment was more common in APS patients compared to age-, gender- and cardiovascular risk factor-matched controls; within one year of follow-up, progression of cognitive impairment occurred in ∼21% of APS patients (13/60), 12 of whom had suffered stroke.^29^ Higher risk of haemorrhagic transformation post APS-associated stroke regardless of thrombolytic therapy was also reported; APS patients spent longer periods in hospital and had a greater risk of mortality despite being significantly younger compared to non-APS stroke patients.^30^

Here, we investigated the impact of aPL in a vascular event setting such as stroke, focusing on the critical period following clot dissolution and brain reperfusion by employing a non-thrombotic outcome. Our study highlights an additional pathogenic effect of aPL that goes beyond lowering the threshold for arterial thrombosis and describes a mechanism for enhanced tissue injury during the reperfusion phase, which may account for the worse prognosis proposed in APS-associated stroke. Patients may benefit from close monitoring, particularly if thrombolysis is given, and incorporating readouts for disability and cognitive impairment during long-term follow up.

We propose that aPL, free to enter the brain following BBB breakdown, impact Akt-mediated homeostatic mechanisms critical to cell survival during reperfusion. Analysis of limited material from our *in vivo* model may indicate a role for the tumour suppressor PTEN in aPL-mediated Akt inhibition. This was a surprising finding as PTEN acts upstream of Akt to prevent activation of phosphoinositide 3-kinase (PI3K)-Akt signalling and may indicate the presence of an inhibitory complex composed of Akt, PTEN and other PI3K signalling components recruited to sites near the plasma membrane.

Another unexpected finding was that, following stroke, renal p-Akt levels were elevated in APS-IgG compared to HC-IgG animals, an observation that agrees with an elegant study for APS nephropathy.^24^ In the absence of stroke, p-Akt levels did not differ between APS-IgG and HC-IgG kidney or brain. We speculate that, similar to the ‘two-hit’ model for thrombotic APS, the presence of circulating aPL combined with the haemodynamic impact of MCA occlusion influences the renal system in our model and drives renal Akt activation.

Both the APS nephropathy study and a second *in vivo* thrombosis model study implicate dysregulation of Akt and its signalling mediators in endothelium, but in opposing ways – suggesting activation^24^ and inhibition^37^ respectively. A key difference between these two studies was the use of exogenous VEGF (a potent Akt inducer) in the latter,^37^ suggesting that aPL may prevent Akt activation following biomechanical challenge. Harnessing this information, we sought to understand the premise of aPL-mediated Akt dysregulation in the presence of an Akt-activating stimulus such as hypoxia-reperfusion. We focused on endothelium, initially showing reduced p-eNOS levels (a downstream Akt target) in brain and carotid artery specimens from APS-IgG animals. A series of *in vitro* experiments corroborated our *in vivo* findings, whereby aPL repressed Akt activation following *in vitro* hypoxia-reperfusion. We utilized two sources of endothelial cells: HUVEC treated with APS-IgG (the most common *in vitro* model for aPL-endothelial studies) and ECFC isolated from patients.

ECFC, considered to originate from true circulating endothelial ‘progenitors’, are capable of clonal expansion and vascular repair, and appear to be ‘biologically primed’ even after long term *in vitro* culture. This precious source of patient endothelium has offered mechanistic insight into endothelial dysfunction across a range of diseases,^31^ including rheumatoid arthritis where ECFC were shown to be hyperproliferative, proangiogenic and sensitized to TNFα stimulation.^39^ Here, we utilise APS ECFC for the first time to demonstrate that both functional (cell death) and molecular (Akt) outcomes are negatively impacted under hypoxia-reperfusion, mirroring our *in vivo* observations and underlining the utility of ECFC for the exploration of endothelial dysfunction in APS.

Our study has some limitations. Our model follows a standard approach of passive transfer of aPL *in vivo* to study APS pathogenesis (‘acute’ APS models). ‘Chronic’ APS models involving immunisation with cardiolipin or β2GPI, are less common but mimic observations from acute models. We used a small number of animals for our *in vivo* model and a moderate number of patients (purified IgG and ECFC) to confirm our findings. *In vivo* experiments employed pooled IgG isolated from two APS and four HC donors. For comparison, our previous studies^9,13,21^ and those by other groups^11,12,48^ also tested a small number of individual APS-IgG or HC-IgG samples in animal models, ranging from 1-6 samples per group. We previously used pooled HC-IgG *in vivo*^9,13^ and pooled APS-IgG and HC-IgG *in vitro* with downstream individual sample validation.^22,49^ Another limitation is that we did not explore reversal of the Akt response with commercially available pan-Akt activators as they are unlikely to be employed in real world clinical practice.

Despite these limitations, we demonstrate consistency across our findings and present novel *in vivo* and *ex vivo* ECFC-based models for oxygen deprivation-reperfusion injury in APS. Future work will aim to establish how aPL-mediated functional and molecular disturbances can be therapeutically reversed. We previously reported on the pro-survival effect of HCQ in an *in vivo* cardiac ischemia-reperfusion model and in cardiomyocytes *in vitro*.^42^ This effect was mediated by ERK and not Akt, but serves as an example of repurposing or reconsidering the benefits of pre-existing drugs already used in autoimmune disease management. Here, using blood-derived ECFC we provide proof-of-principle that HCQ may offer protection against cell death following hypoxia-reperfusion, an effect that is more pronounced in APS compared to HC ECFC. Incorporating HCQ as an adjunct immunomodulatory treatment in APS management is increasingly implemented, and may provide similar benefits to those seen in systemic lupus erythematosus and rheumatoid arthritis where HCQ is proposed to reduce cardiovascular risk.^50^

In conclusion, we translate biological evidence generated from a novel *in vivo* APS stroke model to *in vitro* studies with aPL-treated HUVEC and finally, to APS-derived *ex vivo* EC. We identify an intervenable pathobiological process independent of thrombosis that may inform clinical management of patients with aPL and stroke. Further research is required to establish whether aPL-mediated Akt disturbances can be manipulated therapeutically and the efficacy of immunomodulation towards thrombotic and non-thrombotic pathogenesis in APS.

## Data Availability

Data can be made available following request to the corresponding author.

## Acknowledgments

This manuscript is dedicated to Professor Justin Mason (Imperial College London) and Professor Silvia Pierangeli (University of Texas Medical Branch) who passed away prematurely. We are sincerely grateful to Mrs Valerie Taylor (University College London) for sharing her expertise and supporting the *in vivo* part of this study. Infrastructure support for this research was provided by the NIHR Imperial Biomedical Research Centre (BRC).

---

**Supplementary Table 1.** List of antibodies.

|  | Target Species | Host | Dilution | Supplier | Catalogue number |
| --- | --- | --- | --- | --- | --- |
| **Immunoblot, primary antibody** |  |  |  |  |  |
| Cleaved caspase 3 | Human/Rodent | Rabbit | 1:1000 | Cell Signaling Technology (CST) |  |
| p-Akt S473 | Human/Rodent | Rabbit | 1:1000 | CST | 4070 |
| p-Akt T308 | Human/Rodent | Rabbit | 1:1000 | CST | 13038 |
| p-Bad S136 | Human/Rodent | Rabbit | 1:1000 | CST | 4366/5286 |
| p-eNOS S1177 | Human/Bovine/Pig | Rabbit | 1:1000 | CST | 9571 |
| p-S6RP | Human/Rodent | Rabbit | 1:5000 | CST | 4858 |
| Akt | Human/Rodent | Rabbit | 1:5000 | CST | 4685 |
| Akt | Human/Rodent | Mouse | 1:5000 | CST | 2920 |
| β-actin | Human/Rodent | Mouse | 1:10000 | CST | 3700 |
| Bad | Human/Rodent | Rabbit | 1:1000 | CST | 9292 |
| GAPDH | Human/Rodent | Rabbit | 1:10000 | CST | 2118 |
| PP2AC | Human/Rodent | Rabbit | 1:1000 | CST | 2038 |
| PTEN | Human/Rodent | Rabbit | 1:1000 | CST | 9188 |
| np-PTEN | Human/Rodent | Rabbit | 1:1000 | CST | 7960 |
| S6RP | Human/Rodent | Rabbit | 1:5000 | CST | 2217 |
| vinculin | Human/Rodent | Rabbit | 1:10000 | CST | 13901 |
| **Immunoprecipitation, capture antibody** |  |  |  |  |  |
| Akt | Human | Rabbit | 1:40 | CST | 4685 |
| **Immunoblot, secondary/detection antibody** |  |  |  |  |  |
| Anti-rabbit IgG, horseradish peroxidase | Rabbit | Swine | 1:5000 | Dako | P039901 |
| Anti-mouse IgG, horseradish peroxidase | Mouse | Goat | 1:5000 | Dako | P044701 |
| Anti-rabbit IgG, DyLight™ 800-4x PEG | Rabbit | Goat | 1:5000 | ThermoFisher | SA5-35571 |
| Anti-mouse IgG, AF680 | Mouse | Goat | 1:10000 | ThermoFisher | A32729 |
| Anti-rabbit IgG, conformation specific | Rabbit | Mouse | 1:2000 | CST | 5127 |
| **ECFC purity, flow cytometry** |  |  |  |  |  |
| CD144 PE | Human | Mouse | 10μL | EBioscience | 12-1449-82 |
| CD146 BV421 | Human | Mouse | 5μL | Biolegend | 361004 |
| CD31 FITC | Human | Mouse | 5μL | Biolegend | 303104 |
| CD45 BV510 | Human | Mouse | 5μL | Biolegend | 368526 |
| CD14 APC | Human | Mouse | 5μL | Biolegend | 301808 |
| Zombie NIR™ Fixable Viability Dye | - | - | 1:375 | Biolegend | 423106 |

**Supplementary Figure 1:**
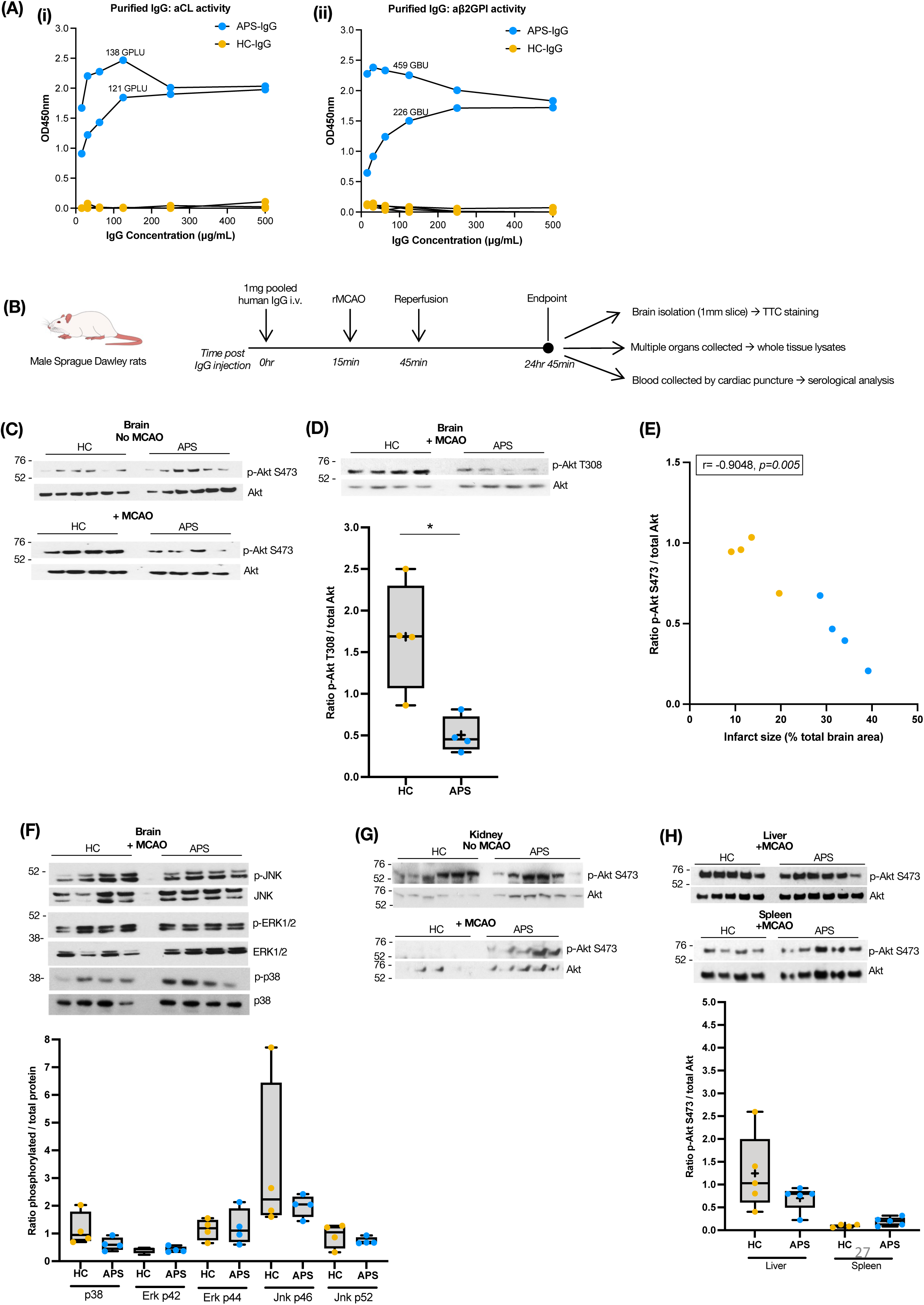
**(A)** Purified APS- and HC-IgG used in the in vivo model was tested for aCL and ab2GPI activity at up to 500ug/mL. ELISA results are shown as net optical density units (OD450nm). Using in-house calibrators with pre-defined activity units,^33^ both APS-IgG preparations were strongly positive (activity units provided for 100ug/mL). HC-IgG were negative (n=2 APS, n=4 HC). **(B)** Schematic of APS *in vivo* stroke model procedures. **(C)** p-Akt S473 blots for individual brain lysates (n=4 per MCAO group, n=6 per no-MCAO group). Normalised to total Akt. Quantification is shown in Figure 2A. **(D)** p-Akt T308 levels in MCAO rat brain revealed lower levels in APS compared to HC animals (n=4 per group). Normalised to total Akt. Results are shown as median ± interquartile range. **(E)** p-Akt levels negatively correlate with infarct size (n=4 per group). *p<0.05. **(F)** p38, ERK1/2 (p44/p42) and JNK1/2 (p54/p46) in MCAO rat brain were similar in APS and HC animals (n=4 per group). Normalised to total p38, ERK1/2, JNK1/2 respectively. Results are shown as median ± interquartile range. **(G)** p-Akt S473 blots for individual kidney lysates (n=5 per APS MCAO, n=4 HC MCAO, n=6 APS and HC no-MCAO). Normalised to total Akt. Quantification is shown in Figure 2H. **(H)** p-Akt S473 was assessed in the liver and spleen of MCAO animals with no apparent differences observed (all n=5 except HC spleen, n=4). Normalised to total Akt. Results are shown as median ± interquartile range.

**Supplementary Figure 2:**
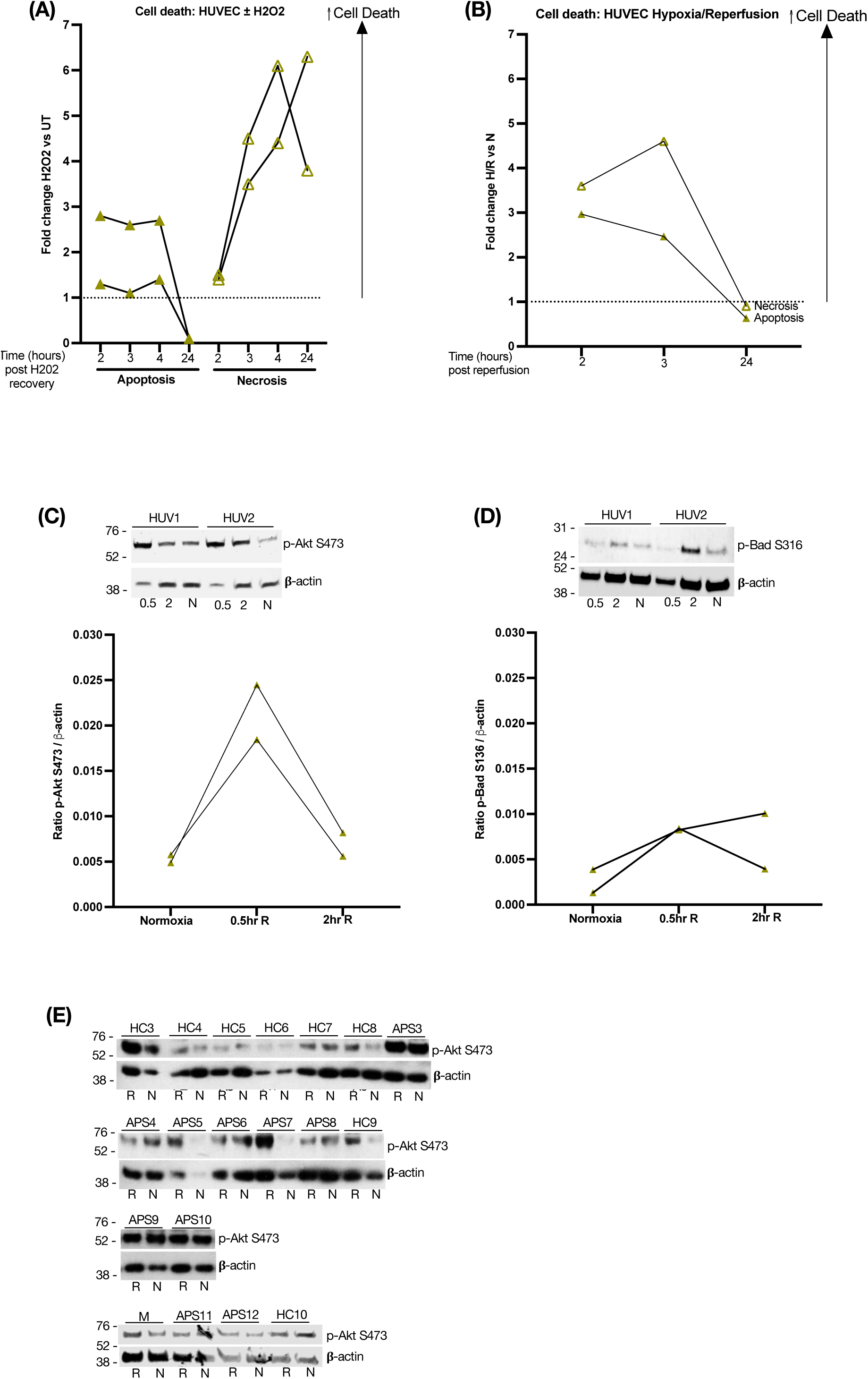
**(A)** As a positive control for the real-time apoptosis/necrosis assay, HUVEC under normoxic conditions were treated for 2hr with 100μM H_2_O_2_ followed by washout/recovery for 24hr. H_2_O_2_ induced both necrosis and apoptosis. Results are shown at 2hr, 3hr and 24hr post-H_2_O_2_ removal and expressed as fold change of apoptosis/necrosis between H_2_O_2_ and untreated HUVEC (n=2 individual HUVEC pools). **(B)** Enhanced cell death in HUVEC subjected to simulated hypoxia-reperfusion *in vitro*, shown at 2hr, 3hr and 24hr post reperfusion. Results are expressed as fold change of apoptosis/necrosis between matched reperfused and normoxic cultures. **(C)** Levels of p-Akt in HUVEC tested at 0.5hr, 2hr reperfusion (0.5R, 2R) and normoxia (N). p-Akt is higher following hypoxia/reperfusion compared to normoxia. Normalised to β-actin. **(D)** Levels of p-Bad in HUVEC tested at 0.5hr, 2hr reperfusion (0.5R, 2R) and normoxia (N). p-Bad is higher following hypoxia-reperfusion compared to normoxia. Normalised to β-actin. **(E)** p-Akt S473 and β-actin blots for individual APS-IgG and HC-IgG treated HUVEC cultures (n=10 APS-IgG, n=8 HC-IgG shown here). M=media alone, untreated. Lysates were collected at 2hr reperfusion (R) or under normoxia (N). Blots from two additional IgG samples per group and total quantification is shown in Figure 3D.

**Supplementary Figure 3:**
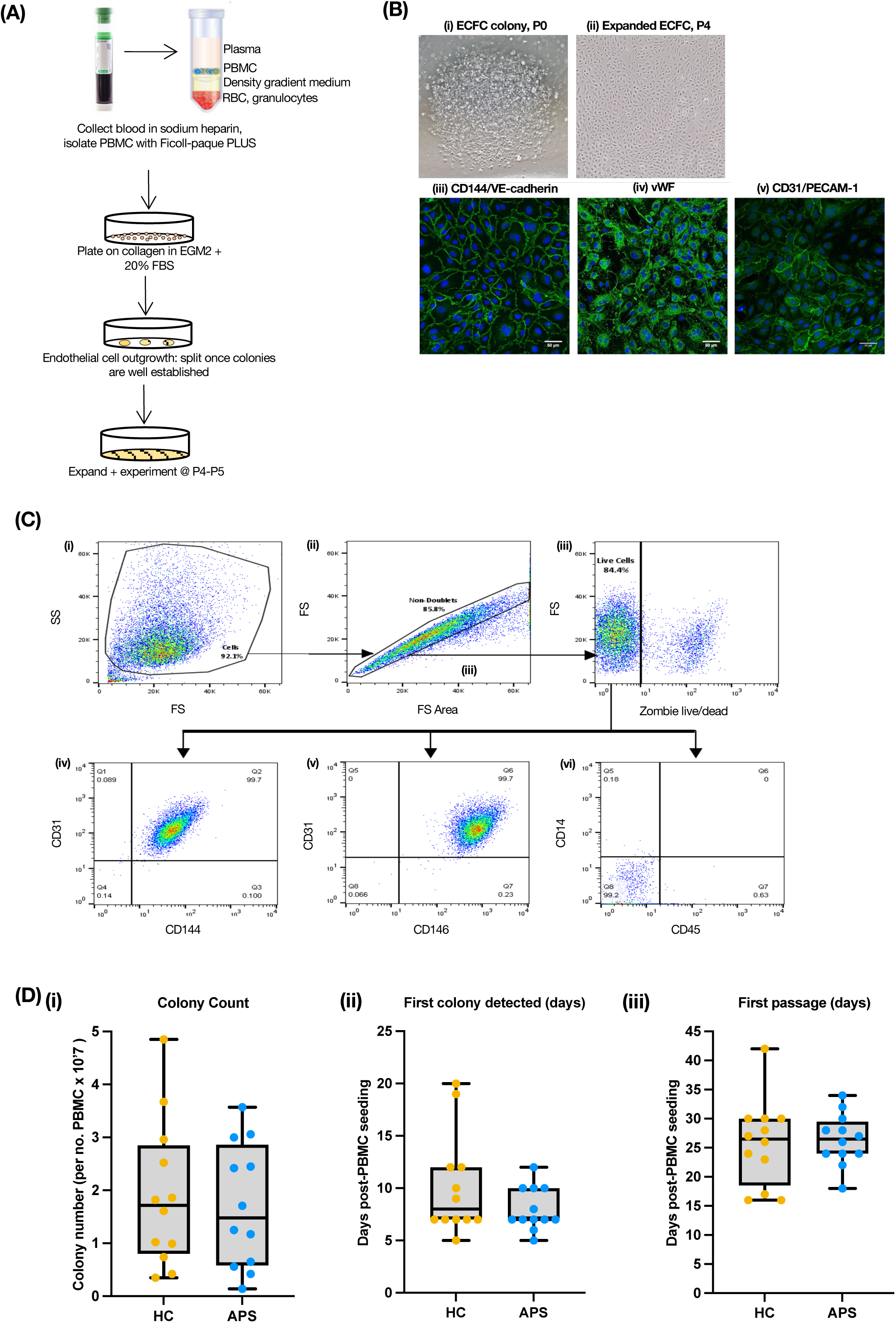
**(A)** Schematic of ECFC isolation. PBMC are plated on collagen coated cell culture vessels and grown in endothelial growth medium-2 (EGM2) supplemented with 20% FBS. Endothelial colonies usually appear within 7-21 days post PBMC seeding and passaged further once well established. **(B)** (i) ECFC colony detected at day 10 post PBMC isolation (left image) and (ii) established ECFC at passage 4 (4x magnification). Representative immunofluorescence images of ECFC at passage 4, stained for canonical endothelial markers (iii) CD144; (iv) von Willebrand factor (vWF); and (v) CD31. **(C)** Flow cytometry gating strategy to characterize endothelial identity of ECFC at passage 4. (i) Forward/side scatter (FS/SS) gating on all cells. (ii) Doublets are excluded to capture only single cells. (iii) Dead cells stained with Zombie dye are excluded. (iv-v) ECFC are CD31/CD144/CD146 positive and (vi) CD45/CD14 negative. **(D)** ECFC generation parameters as recommended by international standardisation guidelines for ECFC isolation.^40^ (i) Colony count: total number of colonies detected, normalised to number of PBMC seeded (x10^7^). (ii) First colony detected: expressed in number of days post PBMC seeding. (iii) First passage: number of days post PBMC seeding where ECFC colonies were passaged from P0 to P1. Results are shown as median ± interquartile range. No significant differences were found.

**Supplementary Figure 4:**
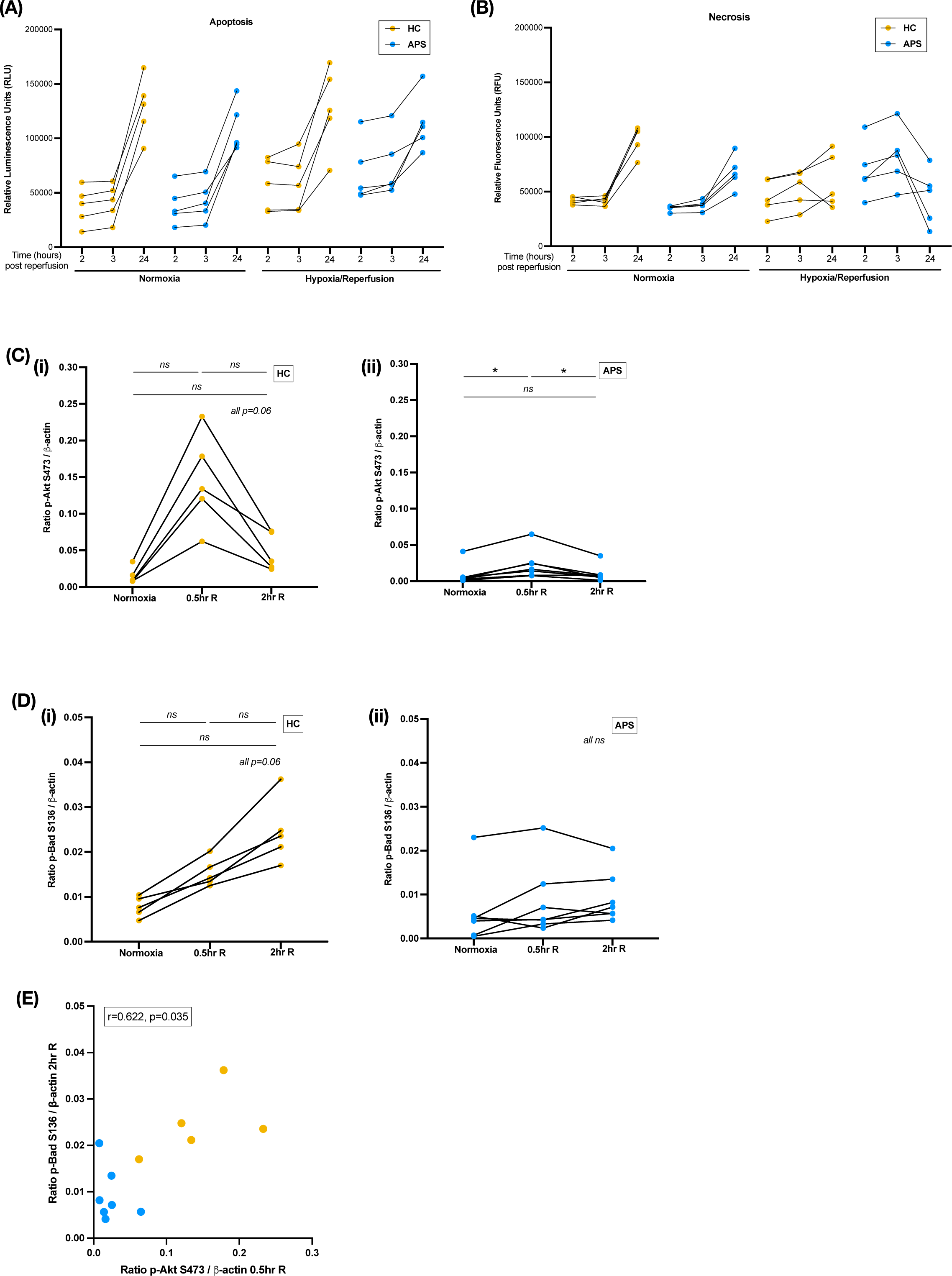
**(A-B)** Representative *(A)* luminescence (apoptosis) and *(B)* fluorescence (necrosis) traces for APS and HC ECFC measured at 2hr, 3hr and 24hr reperfusion or normoxia (n=5 per group). The average of n=3 replicate wells per sample per condition is plotted. **(C)** Levels of p-Akt in (i) HC ECFC and (ii) APS ECFC tested at 0.5hr, 2hr reperfusion (0.5R, 2R) and normoxia (N). Normalised to β-actin. *p<0.05; *ns*, non-significant. **(D)** Levels of p-Bad in (i) HC ECFC and (ii) APS ECFC tested at 0.5hr, 2hr reperfusion (0.5R, 2R) and normoxia (N). Normalised to β-actin. *p<0.05; *ns*, non-significant. **(E)** p-Akt levels at 0.5hr (x-axis) correlate with p-Bad levels at 2hr reperfusion (y-axis).

**Supplementary Figure 5:**
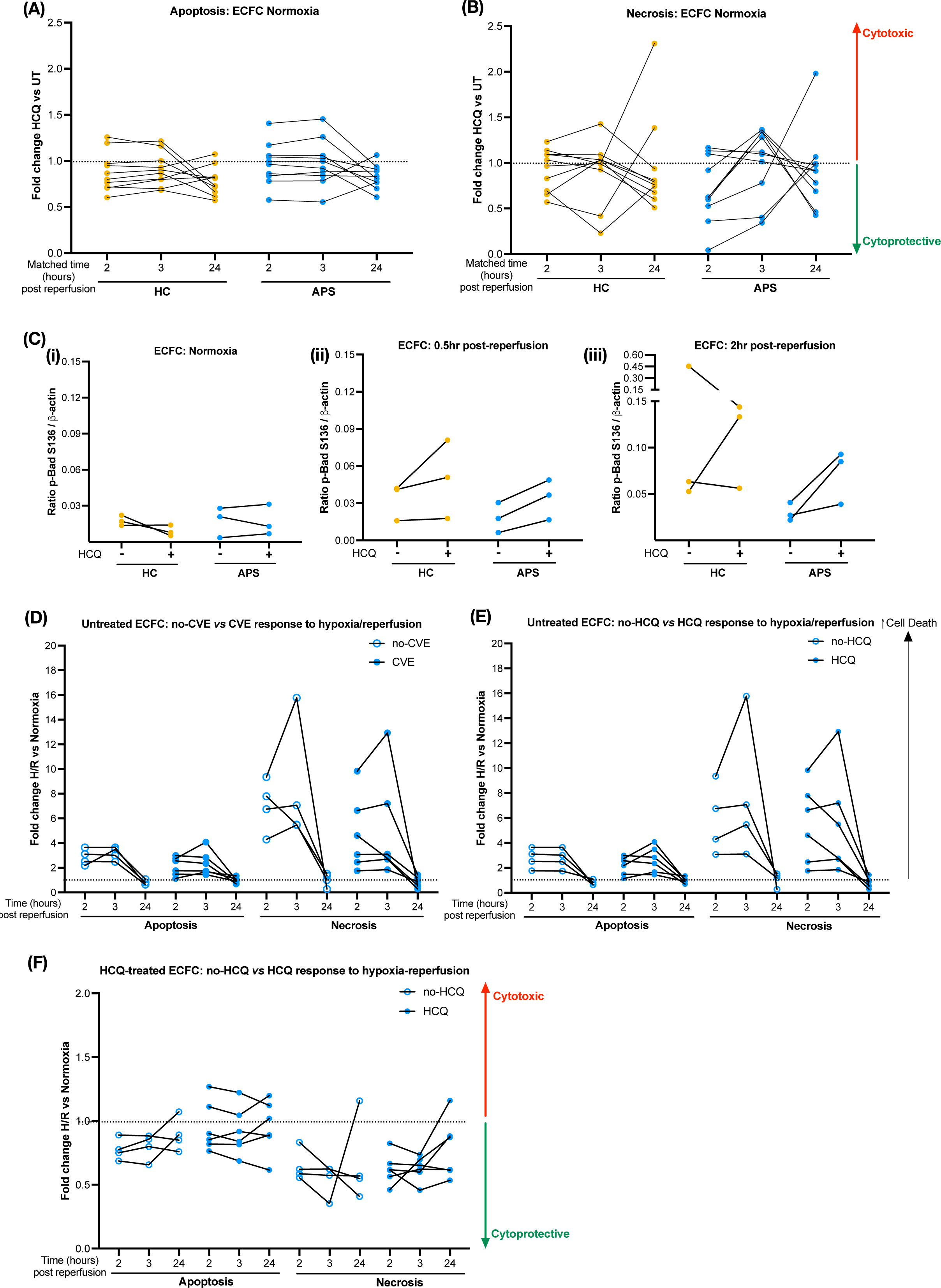
**(A-B)** Variable efffect of hydroxychloroquine (HCQ) on (A) apoptosis and (B) necrosis in ECFC cultured under normoxic conditions. **(C)** p-Bad levels in HC and APS ECFC with or without HCQ treatment under (i) normoxia and following (ii) 0.5hr or (iii) 2hr reperfusion. **(D)** Comparison of cell death responses in ECFC from APS donors with a previous history of cardiovascular events (CVE) *vs* those without (no-CVE). **(E)** Comparison of cell death responses in ECFC from APS donors who received HCQ compared to those not receiving HCQ at the time of ECFC isolation. **(F)** Comparison of cell death responses following exogenous HCQ treatment, between ECFC from APS donors who were on HCQ compared to those not on HCQ at the time of ECFC isolation. **(D-F)** are expressed as fold change of cell death rates under hypoxia-reperfusion (H/R) *vs* normoxia.

